# Scientific Evidence of Prostate Cancer Progression Outcomes in Transgender Females after Hormone Replacement Therapy-Scoping Review Protocol

**DOI:** 10.1101/2024.02.08.24302459

**Authors:** Brenna McAllister, Mylan Panteah, Emily Nelson, Britta Petersen, Katie Hoskins, Sherli Koshy-Chenthittayil, Leslie A. Caromile

## Abstract

Transgender females undergoing hormone replacement therapy (HRT) as a component of the gender affirmation treatment (GAT) commonly retain their prostate, rendering them susceptible to developing prostate cancer (PC). Currently, patients with localized PC receive endocrine therapy (*e.g.,* androgen ablation/castration). Once metastatic, patients undergo standard chemotherapy and/or novel treatment. Unfortunately, many fail to respond completely and develop untreatable, drug-resistant tumors consistent with reprogramming of crucial cell signal transduction pathways that promote tumor growth, invasiveness, and survival. There is no consensus among scientists or physicians on how HRT affects PC treatment options or its related signaling pathways, putting patients at risk for delayed diagnosis. This scoping review aims to analyze and collate the current scientific literature on PC progression in transgender females who have undergone HRT and how PC-initiated oncogenic pathways are impacted by HRT. The review’s findings can potentially inform transgender healthcare and research. This scoping review will follow the Population-Concept-Context methodology for Joanna Briggs Institution Scoping Reviews. Relevant peer-reviewed studies will be identified from the following electronic databases: MEDLINE (PubMed), Embase (Elsevier), CINAHL (EBSCO), and Scopus (Elsevier). Sources of unpublished studies/ grey literature to be searched include bioRxiv (Cold Spring Harbor Laboratory), medRxiv (Cold Spring Harbor Laboratory), and MedNar (Deep Web Technologies). The search strings using keywords such as gender-affirmation treatment, transgender females, and prostate cancer will be conducted using Boolean logic. There will be no limitation on language or date of publication.

## Introduction

Prostate Cancer (PC) is the second most prevalent cancer diagnosed in men in the United States. Early detection is crucial to ensuring successful treatment and survival rates. Risk factors include age, obesity, and ethnicity, but one significant factor is genetic predisposition. A family history of three or more diagnosed individuals significantly increases a patient’s risk of developing PC^1^. If caught early, PC is highly treatable, with a 5-year survival rate of 99%. However, the absence of discernible symptoms during the initial stages often results in delayed diagnosis, leading to metastasis and lowering the survival rate to 30% over 5 years^2^. Early detection of PC is crucial for successful treatment. Promoting screening and developing advanced treatment options can significantly reduce the incidence and severity of this disease and improve the quality of life. Therefore, a comprehensive approach that prioritizes both prevention and treatment strategies is necessary for mitigating the impact of PC.

Testosterone (T) is a critical player in the normal development of the prostate and PC. T diffuses into prostate epithelial cells and converts to dihydrotestosterone (DHT) by the enzyme SRD5A^3^. DHT or T binds androgen receptors (AR) in the cytoplasm, translocating them to the nucleus to regulate gene expression involved in prostate growth^3^. In initial stages, PC can be managed through watchful waiting, surgery, androgen deprivation therapy (ADT), or radiation therapy (docetaxel, enzalutamide, luteinizing hormone-releasing hormone (LHRH) agonists or antagonists)^1,4^. Although PC tumors initially respond to ADT, most men eventually develop a progressive disease resistant to treatment, androgen-independent (castration-resistant prostate cancer [CRPC]), and typically metastatic.

Until recently, the dialogue around PC has focused exclusively on cisgender men. However, the conversation must evolve to include the transgender community, specifically transgender females or individuals who identify as female and not with their birth-assigned male sex. During gender transition from male to female, patients may choose to undergo gender affirmation treatment (GAT), which can involve surgery combined with hormone replacement therapy (HRT) to suppress secondary male sex characteristics^5,6^. HRTs, such as spironolactone and cyproterone acetate, decrease androgen levels, block the AR and supplement estrogen (estradiol)^5^. Other HRTs, such as dutasteride and finasteride, prevent the SRD5A enzyme from converting T to DHT^7,8^. During gender confirmation surgery, the prostate is typically not removed to prevent complications, such as urinary and stress incontinence, leaving transgender females still at risk for developing PC^5,9^.

In some patients, the reduction of androgen levels in transgender females may aid in PC prevention. However, this is not always the case, as estrogen therapy has the potential to augment a patient’s pre-existing risks^10,11^. For example, estrogen and progesterone, rather than T or DHT, can activate the AR containing a common T878A mutation^12^, leading to modulation of NF-kB signaling, MAPK pathway, JAK/STAT pathway, and WNT pathways^13^. Additionally, the supplementation of estrogen in transgender females can activate the alpha or beta subtypes of the estrogen receptor, also leading to downstream changes in oncogenic signaling cascades^14,15^. Moreover, depending on the age the patient decides to begin their transition, HRT can begin anywhere from under 18 years to over 50 years old. While there is no consensus on the role age plays, it is an important factor to consider that has the potential to influence a patient’s response to HRT^16,17^.

Previous reviews on PC in transgender females have focused on clinical case reports, clinical screening guidelines, and biostatistical data. In addition, recent studies have investigated the effects of HRT on oncogenic signaling pathways. However, to our knowledge, no review has connected these two concepts. Therefore, this scoping review aims to analyze and collate the current scientific literature on PC progression in transgender females who have undergone HRT and how PC-initiated oncogenic pathways are impacted by HRT.

## Methods

The stages of the review include (1) identifying the research questions, (2) identifying the relevant studies, (3) study selection, (4) charting the data, (5) collating, summarizing, and reporting the results, and (6) consultation.

### (1) Identifying the research question

Based on the Population, Concept, Context (PCC) framework put forward by JBI^18^, the research question for this scoping review is “What are the reported outcomes of PC progression in the transgender female community that has undergone HRT?” The sub-question is: “What molecular mechanisms or pathways have been altered because of HRT in the transgender female community?” **Table 1** shows the PCC regarding our review.

**Table 1:** Population-Concept-Context Methodology.

|  |  |
| --- | --- |
| **Population** | transgender females who have undergone the transition from male to female, or who have undergone HRT, or transgender females who have been clinically diagnosed with PC, or who have a family history of PC. |
| **Concept** | PC development/treatment/detection in transgender females exclusively unless presented in comparison to PC in cismen |
| **Context** | healthcare and research settings worldwide |

### (2) Identifying the relevant studies

The proposed scoping review will follow the JBI methodology for scoping reviews^18^. The final manuscript will also follow the Preferred Reporting Items for Systematic Reviews and Meta-Analyses Extension for Scoping Reviews (PRISMA-ScR)^19^. The search strategy will aim to locate both published and unpublished studies. An initial limited search of PubMed was undertaken while consulting with librarians from UConn Health and Touro University to identify articles on the topic. The text words contained in the titles and abstracts of relevant articles and the index terms used to describe the articles were used to develop a full search strategy for PubMed (**see Appendix 1**). The search strategy, including all identified keywords and index terms, will be adapted for each included database and/or information source. The reference list of all included sources of evidence will be screened for additional studies. There are no restrictions on the date or time of publication or study. The databases to be searched include MEDLINE (PubMed), Embase (Elsevier), CINAHL (EBSCO), and Scopus (Elsevier). Sources of unpublished studies/ grey literature to be searched include bioRxiv (Cold Spring Harbor Laboratory), medRxiv (Cold Spring Harbor Laboratory), and MedNar (Deep Web Technologies).

### (3) Selection of studies

Following the search, all identified citations will be collated and uploaded into Covidence (Veritas Health Innovation Ltd, Melbourne, Australia), and duplicates will be removed by the reviewers. Following a pilot test, titles and abstracts will then be screened by three or more independent reviewers for assessment against the inclusion criteria for the review. Potentially relevant sources will be retrieved in full. Three or more independent reviewers will thoroughly assess the full text of selected citations against the inclusion criteria. Reasons for excluding sources of evidence in full text that do not meet the inclusion criteria will be recorded and reported in the scoping review. Any disagreements that arise between the reviewers at each stage of the selection process will be resolved through discussion or with an additional reviewer/s. The search results and the study inclusion process will be reported in full in the final scoping review and presented in a Preferred Reporting Items for Systematic Reviews and Meta-analyses extension for scoping review (PRISMA-ScR) flow diagram^19^.

This scoping review will consider both experimental and quasi-experimental study designs, including randomized controlled trials, non-randomized controlled trials, and interrupted time-series studies. In addition, analytical observational studies, including prospective and retrospective cohort studies, case-control studies, and analytical cross-sectional studies, will be considered for inclusion. This review will also consider descriptive observational study designs, including case series, individual case reports, and descriptive cross-sectional studies for inclusion. In addition, systematic reviews that meet the inclusion criteria will also be considered, depending on the research question. The review will include animal and cell culture models. Text and opinion papers will be excluded from this scoping review because of potential bias towards the transgender community. No ongoing clinical trials will be included in this review.

In this review, we will be evaluating the current literature on PC progression and outcomes in transgender females who have undergone HRT and highlighting links between HRT and known oncogenic molecular mechanisms involved in PC progression.

### The eligibility criteria for this study will be as follows

#### Inclusion Criteria for This Study

- Articles focused on transgender females who have undergone HRT.
- Articles focused on transgender females with a family history of PC.
- Articles focused on transgender females clinically diagnosed with PC.
- Articles focused on molecular mechanisms involved in PC progression in transgender females.
- Articles published worldwide.

#### Exclusion Criteria for This Study

- Articles exclusively focused on PC in cis males.
- Articles exclusively focused on cancers other than PC unless containing information about PC in transgender females.
- Biased opinion papers.

**Figure 1:**
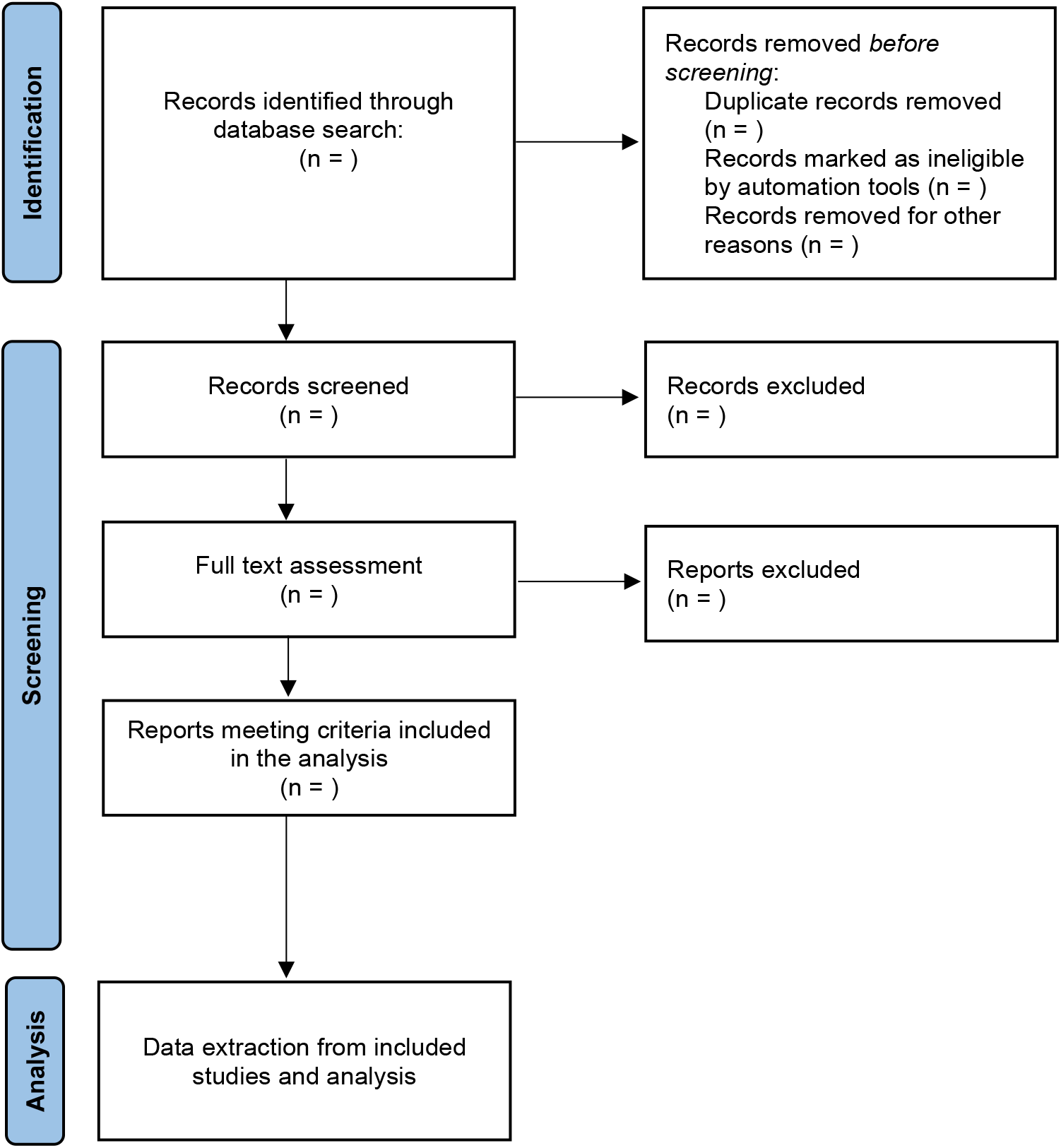
PRISMA Flow Diagram. Selection of studies according to PRISMA-ScR protocol. *Adapted From:* Page MJ, McKenzie JE, Bossuyt PM, Boutron I, Hoffmann TC, Mulrow CD, et al. The PRISMA 2020 statement: an updated guideline for reporting systematic reviews. BMJ 2021;372:n71. doi: 10.1136/bmj.n71

### (4) Charting the data

Data will be extracted from papers included in the scoping review by three or more independent reviewers using a data extraction tool developed by the reviewers. A draft extraction form is provided (**see Appendix 2**). The draft data extraction tool will be modified and revised as necessary while extracting data from each included evidence source. Modifications will be detailed in the scoping review. Any disagreements that arise between the reviewers will be resolved through discussion or with an additional reviewer/s. The key information that reviewers have been chosen to chart:

From these sources, this scoping review will extract data on the GAT received (age, treatment duration, types of HRT), the methodology used for PC detection and treatment, rates of PC in transgender females, molecular pathways altered by HRT and any key findings relevant to our objective and review question.

### (5) Collating, summarizing, and reporting the results

As this is a scoping review, the results of the data extraction will provide a summary and collation of the PC progression and outcomes in transgender females who have undergone HRT. Due to the possible heterogeneity of the studies, we hope to use either a narrative analysis and/or descriptive figures/tables to depict the results. The final manuscript will follow the PRISMA-ScR format extension.

### (6) Consultation

The results will be disseminated using paper publications and presentations at national conferences such as ASCB (American Society for Cell Biology) and local conferences. The link to the open-source paper will be publicly available.

## Discussion

Prostate cancer is the fifth leading cause of cancer-related deaths in men worldwide and has typically only been studied in cis men^20,21^. Although transgender women are still capable of developing PC, until recently, they were excluded from research on primary care. Contributing to this disparity in patient care, discrimination and restricted access to transgender health care make it difficult for patients to create a prevention plan, leaving them vulnerable to delayed diagnosis^10,22–27^.

Current reviews have focused on advising the medical community on patient care or screening guidelines. This scoping review aims to provide information on the relationship between PC progression in transgender women and oncogenic signaling pathways impacted by HRT to recommend a direction for future transgender research and highlight the importance of secure transgender healthcare. The main strength of this scoping review is the new perspective it offers by connecting these two topics that have only been viewed separately. It is important to note there are several limitations of this scoping review. The quality and accessibility of healthcare for transgender individuals varies significantly worldwide due to several barriers, including cultural, socioeconomic, and political; the results and suggested future directions from this scoping review may be applied in global healthcare and research settings at varying rates. Furthermore, transgender women may experience emotions of gender dysphoria or distress related to the misalignment of their sex and gender identity when discussing prostate health or screening procedures^28^. Additionally, the scant number of documented cases resulting from mistrust between the transgender population and the medical establishment has resulted in a paucity of published scientific evidence on PC in the transgender community ^29–33^. Although this scoping review will consider worldwide publications, certain countries may have more published studies on this topic than others. Despite these limitations, this scoping review has the potential to raise awareness about the transgender community and assist future research in their goals to enhance PC prevention and treatment for all individuals afflicted by the disease.

## Data Availability

Data will be made publicly available when the study is completed and published.

## Acknowledgements

We thank Marissa Iverson, the research and instructional services librarian at UConn Health Sciences Library, and Megan De Armond, librarian at Touro University Nevada, for their help with our initial search.

