## Appendix 1 for "Scientific Evidence of Prostate Cancer Progression Outcomes in Transgender Females after Hormone Replacement Therapy-Scoping Review Protocol"

***Appendix 1: Search Strategy***

Initial Search: MEDLINE (PubMed)
Date of Search: 9-27-2023

| | | |
|---|---|---|
| #1 | "cancer of prostate"[tiab] OR "cancer of the prostate"[tiab] OR "familial prostate cancer*"[tiab] OR "genitourinary malignanc*"[tiab] OR "hereditary prostate cancer"[tiab] OR "malignant prostate tumo*"[tiab] OR "malignant prostatic tumo*"[tiab] OR "malignant tumo*"[tiab] OR "prostate cancer*"[tiab] OR "prostate cancer, familial" [Supplementary Concept] OR "prostate carcinogenesis"[tiab] OR "prostate gland cancer*"[tiab] OR "prostate malignanc*"[tiab] OR "prostate malignant tumo*"[tiab] OR "prostate neoplas*"[tiab] OR "prostatic cancer*"[tiab] OR "prostatic carcinogenesis"[tiab] OR "prostatic neoplas*"[tiab] OR "prostatic neoplasms"[Mesh] OR "sex hormone related cancer*"[tiab] | 257,607 |
| #2 | "birth-assigned male*"[tiab] OR "cross-sex hormone*"[tiab] OR "female gender identity"[tiab] OR "feminis*"[tiab] OR "feminization"[Mesh] OR "gender dysphoria"[Mesh] OR "gender dysphoria"[tiab] OR "gender identity"[Mesh] OR "gender identity"[tiab] OR "gender minority adult*"[tiab] OR "gender non-conforming"[tiab] OR "gender reassignment"[tiab] OR "male to female"[tiab] OR "natal male*"[tiab] OR "sex reassignment procedures"[Mesh] OR "sex reassignment"[tiab] OR "sexual and gender minorities"[Mesh] OR "trans people"[tiab] OR "trans person*"[tiab] OR "transfeminine"[tiab] OR "transgender persons"[Mesh] OR "transgender*"[tiab] OR "transpeople*"[tiab] OR "transperson*"[tiab] OR "transsexual*"[tiab] OR "transsexualism"[Mesh] OR "transwomen"[tiab] OR "two spirit person*"[tiab] | 65,511 |
| #3 | "5-alpha reductase inhibitors"[Mesh] OR "5-alpha reductase inhibitor*"[tiab] OR "androgen antagonists"[Mesh] OR "androgen receptor antagonist*"[tiab] OR "androgen receptor antagonists"[Mesh] OR "Receptors, Androgen"[Mesh] OR "androgen receptor*"[tiab] OR "androgen suppression"[tiab] OR "antiandrogen"[tiab] OR "estradiol/therapeutic use"[Mesh] OR "Receptors, Estradiol"[Mesh] OR "estrogen therapy"[tiab] OR "estrogens/therapeutic use"[Mesh] OR "gender-affirming hormon*"[tiab] OR "gonadotropin-releasing hormone*"[tiab] OR "gonadotropin-releasing hormone"[Mesh] OR "hormone replacement therap*"[tiab] OR "hormone replacement therapy"[Mesh] OR "hormone substitution"[tiab] OR "reductase inhibitor*"[tiab] OR "Androcur"[tiab] OR "cyprostat"[tiab] OR "cyproterone acetate"[tiab] OR "Cyproterone Acetate"[Mesh] OR "Avodart"[tiab] OR "GG745"[tiab] OR "GI198745"[tiab] OR "dutasteride"[Mesh] OR "dutasteride"[tiab] OR "Propecia"[tiab] OR "Proscar"[tiab] OR "MK-906"[tiab] OR "MK906"[tiab] OR "finasteride"[Mesh] OR "finasteride"[tiab] OR "Eulexin"[tiab] OR "Niftolid"[tiab] OR "Niftolide"[tiab] OR "Chimax"[tiab] OR "Euflex"[tiab] OR "Drogenil"[tiab] OR "Fluta*"[tiab] OR "Fugerel"[tiab] OR "SCH- | 319,755 |

| | | |
|---|---|---|
| | 13521"[tiab] OR "SCH13521"[tiab] OR "flutamide"[Mesh] OR "flutamide"[tiab] OR "Supprelin"[tiab] OR "Vantas"[tiab] OR "histrelin"[Supplementary Concept] OR "histrelin"[tiab] OR "Aldactone"[tiab] OR "CaroSpir"[tiab] OR "Spiro*"[tiab] OR "Veroshpiron"[tiab] OR "Verospiron"[tiab] OR "Verospirone"[tiab] OR "Spiractin"[tiab] OR "Aquareduct"[tiab] OR "Espironolactona*"[tiab] OR "SC-9420"[tiab] OR "SC9420"[tiab] OR "spironolactone"[Mesh] OR "spironolactone"[tiab] OR "Anandron"[tiab] OR "RU-23908"[tiab] OR "RU23908"[tiab] OR "nilutamide" [Supplementary Concept] OR "estradiol"[tiab] OR "oestradiol"[tiab] OR "Progynova"[tiab] OR "Vivelle"[tiab] OR "Aerodiol"[tiab] OR "Estrace"[tiab] OR "Estraderm"[tiab] OR "Progynon Depot"[tiab] OR "Delestrogen"[tiab] OR "Ovocyclin"[tiab] OR "Alora"[tiab] OR "Climara"[tiab] OR "Delestrogen"[tiab] OR "Divigel"[tiab] OR "Dotti"[tiab] OR "Elestrin"[tiab] OR "Estrace"[tiab] OR "Estrogel"[tiab] OR "Evamist"[tiab] OR "Femring"[tiab] OR "Menostar"[tiab] OR "Minivelle"[tiab] OR "Climara"[tiab] OR "Estradot"[tiab] OR "Oesclim"[tiab] OR "Estradiol"[Mesh] OR "Leuprorelin"[tiab] OR "Enantone"[tiab] OR "Leuprolide Acetate"[tiab] OR "Lupron"[tiab] OR "TAP-144"[tiab] OR "TAP144"[tiab] OR "A-43818"[tiab] OR "A43818"[tiab] OR "Eligard"[tiab] OR "Fensolvi"[tiab] OR "leuprolide"[tiab] OR "Leuprolide"[Mesh] OR "ICI-118630"[tiab] OR "ICI118630"[tiab] OR "Zoladex"[tiab] OR "goserelin"[tiab] OR "Goserelin"[Mesh] | |
| #4 | #1 AND #2 AND #3 | 72 |
