## Appendix 2 for "Scientific Evidence of Prostate Cancer Progression Outcomes in Transgender Females after Hormone Replacement Therapy-Scoping Review Protocol"

***<u>Appendix 2: Data extraction instrument (has been modified from JBI instrument)</u>***

| |
|---|
| Author: |
| Year of Publication: |
| Origin/Country of Origin: |
| Aims/Purpose: |
| Study Population and Sample Size (If applicable): |
| Information about Male to Female Transition (age, duration, types of hormone treatments, etc.): |
| The technique used for Prostate Cancer Detection (If applicable): |
| The technique used for Prostate Cancer Treatment (If applicable): |
| Family history of prostate cancer (if applicable): |
| For comparison papers, information about the control population: |
| Molecular pathways/mechanisms that were altered due to HRT: |
| Key Findings: |
| Other: |
